# Clinical identification of malignant pleural effusions

**DOI:** 10.1101/2020.05.31.20118307

**Authors:** Jianlong Jia, Antonia Marazioti, Apostolos Voulgaridis, Ioannis Psallidas, Anne-Sophie Lamort, Marianthi Iliopoulou, Anthi C. Krontira, Ioannis Lilis, Rachelle Asciak, Nikolaos I. Kanellakis, Najib M. Rahman, Kyriakos Karkoulias, Konstantinos Spiropoulos, Ruonan Liu, Jan-Christian Kaiser, Georgios T. Stathopoulos

**Author notes:** Corresponding author: Georgios T. Stathopoulos, Comprehensive Pneumology Center, Max-Lebsche-Platz 31, 1. OG, Munich, 81377, Germany. These authors contributed equally to this work as senior authors (Jan-Christian Kaiser, Georgios T. Stathopoulos).

## Abstract

**Importance:** Pleural effusions frequently signal disseminated cancer. Diagnostic markers of pleural malignancy at presentation that would assess cancer risk and would streamline diagnostic decisions remain unidentified.

**Objective:** The present study aimed at identifying and validating predictors of malignant pleural effusion at patient presentation.

**DESIGN, SETTING, AND PARTICIPANTS:** A consecutive cohort of 323 patients with pleural effusion (PE) from different etiologies were recruited between 2013-2017 and was retrospectively analyzed. Data included history, chest X-ray, and blood/pleural fluid cell counts and biochemistry. Group comparison, receiver-operator characteristics, unsupervised hierarchical clustering, binary logistic regression, and random forests were used to develop the malignant pleural effusion detection (MAPED) score. MAPED was validated in an independent retrospective UK cohort (*n* = 238).

**Main Outcomes and Measures:** The outcome was diagnostic of pleural effusion in patients, and the clinical and laboratory indicators available of the patient were measured.

**Results:** Five variables showed significant diagnostic power and were incorporated into the 5-point MAPED score. Age > 55 years, effusion size > 50% of the most affected lung field, pleural neutrophil count < 2,500/mm^3^, effusion protein > 3.5 g/dL, and effusion lactate dehydrogenase > 250 U/L, each scoring one point, predicted underlying cancer with the area under curve(AUC) = 0.819 (sensitivity=82%, specificity=74%, *P* < 10^-15^) in the derivation cohort. The AUC and net reclassification improvement (NRI) of MAPED score and cytology were not significantly different. However, the integrated discrimination improvement (IDI) of The MAPED score displayed a slight increment *(P* <0.001). The calibration curves of the cytology model were slightly better than The MAPED score. Decision curve analysis (DCA) indicated that The MAPED score generated net clinical benefit. In the validation dataset, the results were generally consistent with the above findings, with an AUC of 0.723 (sensitivity=76%, specificity=62%, P =3*10-9) for The MAPED score. Interestingly, MAPED correctly identified 33/42(79%) of cytology-negative patients that indeed had cancer. The MAPED score is used to create nomogram so clinicians can predict the probability of malignant pleural effusions.

**Conclusions:** The MAPED score identifies malignant pleural effusions with satisfactory accuracy and can be used complementary to cytology to streamline diagnostic procedures.

## INTRODUCTION

Pleural effusions (PE) are common conditions that annually affect 1.5 million individuals in the US alone^(1)^. PE are caused by involvement of the pleural space by cancer (malignant PE, MPE) or by non-cancerous processes including infection, inflammation, and deranged Starling or oncotic pressures along juxtapleural blood and lymphatic vessels (benign PE, BPE) ^(1)^. Patients with PE face a markedly dichotomous outcome: a diagnosis of MPE portends a median survival of a few months ^(2–4)^, while patients with BPE fare significantly better ^(1)^. While the time and procedures required for a definitive cell- or tissue-based diagnosis of MPE or an etiologic diagnosis of BPE are substantial, a simple model to predict malignancy that would rapidly inform physicians of the probability of cancer is missing. Even cytological examination of three pleural fluid specimens obtained on consecutive days, considered to be the gold standard in MPE diagnosis together with pleural tissue biopsy, is only successful in two-thirds of MPE cases overall ^(2–5)^.

To bridge this gap, we retrospectively evaluated 323 consecutive patients with PE that were admitted to our emergency wards between 2013 and 2017. We collected all clinical, pleural fluid and blood, and chest X-ray data that were available at patient presentation, and performed detailed examination of pleural cells on May-Grünwald-Giemsa-stained cytocentrifugal specimens. MPE-BPE comparisons, receiver-operator characteristics, unsupervised hierarchical clustering, binary logistic regression, and random forests identified five variables that independently predict MPE and that were compiled into the MPE detection (MAPED) model. MAPED displayed area under curve (AUC) = 82% in the derivation cohort and AUC = 72% in a validation cohort of 238 patients with PE from the Oxford Radcliffe Pleural Biobank (ORPB). Interestingly, MAPED performed complementary to cytology.

## METHODS

### Patients

MAPED and ORPB abided by the Helsinki Declaration, were prospectively approved (University of Patras Ethics Committee #22699/21.11.2013 and South Central Oxford A Ethics Committee #15/SC/0186), and all patients gave written informed consent. All 460 adults with a PE that were admitted to the University Hospital of Patras, Greece, between 21/11/2013–21/11/2017, were evaluated. One hundred and thirty seven patients were excluded due to immediate discharge, previous pleural disease/cancer, and/or missing data. We recorded chest X-ray and blood/PE cell, biochemistry, and pH data. Since routine effusion cell counts were done on smears, we additionally counted 400 cells/patient using May-Gruenwald-Giemsa-stained cytocentrifugal specimens (50,000 cells; 300 g; 10 min; 4 ^0^C; Cellspin, Tharmac, Wiesbaden, Germany)^(6)^. PE size was defined on the most affected lung field: 1, ≤ 10%; 2, 11-25%; 3, 26-50%; 4, 51-75%; and 5, > 75%. MPE was diagnosed by positive cytology of any of three consecutive daily effusion samples (50-200mL). Cytology-negative patients were further investigated with thoracoscopy and/or computed tomography-guided biopsy. BPE was diagnosed using established diagnostic criteria (positive pleural fluid smears, cultures, or polymerase chain reaction for common pathogens or Mycobacteria; lymphocytic-predominant exudative effusion with recent tuberculin skin test conversion or conversion within a month after admission; full remission of PE and lung lesions on empiric antibacterial or antituberculous treatment; caseating granulomas in pleural tissue; transthoracic echocardiography-determined ejection fraction < 40% with/without tricuspid regurgitation and/or diastolic dysfunction and/or elevated serum N-terminal pro-B-type natriuretic peptide levels; etc.), according to current guidelines ^(1-4)^.

### Statistics and analyses

Sample size (*n*) was determined using G*Power (https://www.psychologie.hhu.de/arbeitsgruppen/allgemeine-psychologie-und-<u>arbeitspsychologie/gpower</u>)^(7)^. Employing Fischer’s exact test, α and β = 0.05, and 1:1 allocation, *n* = 100-334 was required to detect 20% proportion inequalities between two independent groups, depending on the proportion range (0-100%). Employing Student’s t-test, α and β = 0.05, and 1:1 allocation ratio, *n* = 328 was required to detect effect size *d* = 0.4 between two independent groups. All data were not normally distributed using Kolmogorov-Smirnov test, hence data are given as frequencies or as median (95% confidence interval, 95%CI). Differences between variables were examined using Fischer’s exact, χ^2^, or Kolmogorov-Smirnov tests. Probability (*P*) < 0.05 was considered significant. Bonferroni-corrected *P* was used for multiple comparisons. Receiver-operator characteristics, unsupervised hierarchical clustering, binary logistic regression using backward Waldman elimination, and random forests were done on Project R*^(8)^ and Prism v8.0 (GraphPad, San Diego, CA). Random forests were grown using R* package randomForest (https://cran.r-project.org/web/packages/randomForest/randomForest.pdf) using log-transformed cell counts, lactate dehydrogenase (LDH), and glucose. The top five variables by the criteria of low *P* values and mean decreased accuracy were chosen for MAPED, and cut points were determined on partial dependency plots. MAPED was compared with logistic regression and random forest models including all variables, or only variables with lowest Akaike information criteria (AIC) or top variable importance rank (VIMP), using fifty hold-out resampling repeats of 70:30 train/test data sets to calculate AUC and Brier score/skill. The R package “rms” is employed to plot the fit curves of the observed and predicted values. The R package “PredictABEL” is utilized to calculate the net reclassification improvement (NRI) and the integrated discrimination improvement (IDI), which indicate the discrimination of the prediction model ^(9,10)^. The AIC and the Bayesian Information Criterion (BIC), which demonstrate the calibration of the same model, are also applied ^(11)^. Decision curve analysis (DCA), a measurement of the net clinical benefits, was analyzed using the “Decision Curve” package in R^(12).^ The nomogram is used to construct the scoring system using the “rms” package in R. For each repeat, a confusion test matrix for *P*_MPE_ > 0.5 was used to calculate accuracy, sensitivity, and specificity.

## RESULTS

### Distinctive features of malignant versus benign pleural effusions

The flow graph shows study populations and research project procedures (Figure 1). In total, 323 patients were analyzed in the MAPED study, 189 with BPE and 134 with MPE (Table 1). Most patients with MPE received cytologic diagnoses (*n* = 92), while 52 received tissue-based, and ten both tissue-and cell-based MPE diagnoses. Cytology was used as the reference standard against which all MAPED variables were tested. Out of the 134 patients with MPE, sixty patients had lung cancer (45%), 30 breast cancer (23%), 21 malignant pleural mesothelioma (16%), 10 gynecological malignancies (7%), four gastrointestinal tumors (3%), five hematological malignancies (4%), and four other cancers (3%), proportions that are in accord with other European studies ^(2,4)^. Several differences were identified between patients with BPE and MPE in the 34 different variables examined, using the Bonferroni-adjusted probability threshold of *P* < 0.05/34 = 0.0015 (Figures 2A and 2B, and Table 1). To this end, MPE were more frequently cytology positive and larger in size compared with BPE (Figure 2A). In addition, patients with MPE displayed increased age, PE LDH levels, and PE/blood LDH and protein ratios (Figure 2B). Receiver-operator analyses targeted at MPE identified cytology, PE size, PE neutrophil percentage, and PE-to-blood protein ratio as inputs significantly associated with incipient MPE diagnosis (Figure 2C). Interestingly, PE LDH levels and PE-to-blood LDH and protein ratios represent Light’s criteria used to distinguish exudates from transudates ^(13)^. In summary, comparative analyses identified variables with some stand-alone predictive power of MPE, which was clearly inferior in comparison to cytology.

**Figure 1.**
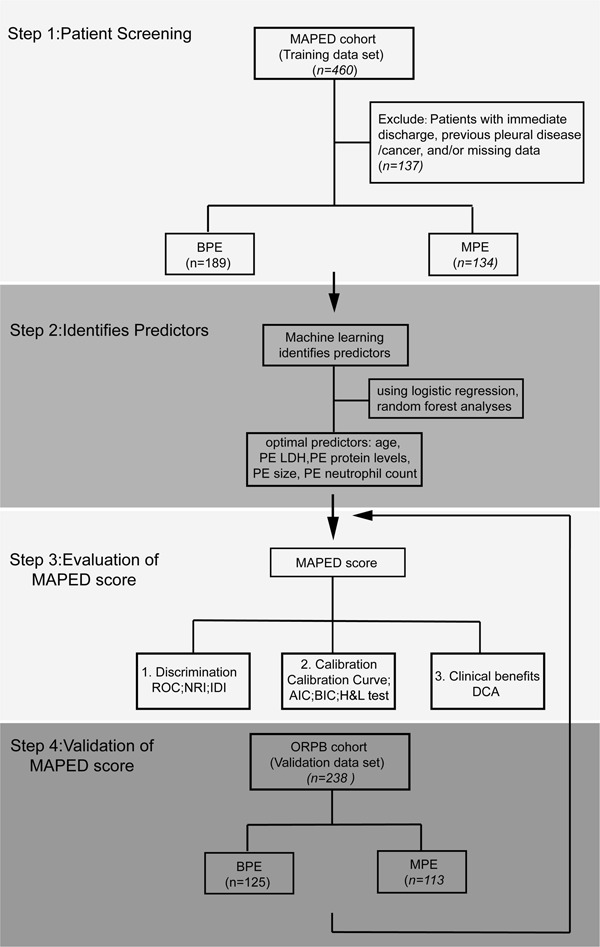
The flow graph shows study populations and research project procedure.

**Figure 2.**
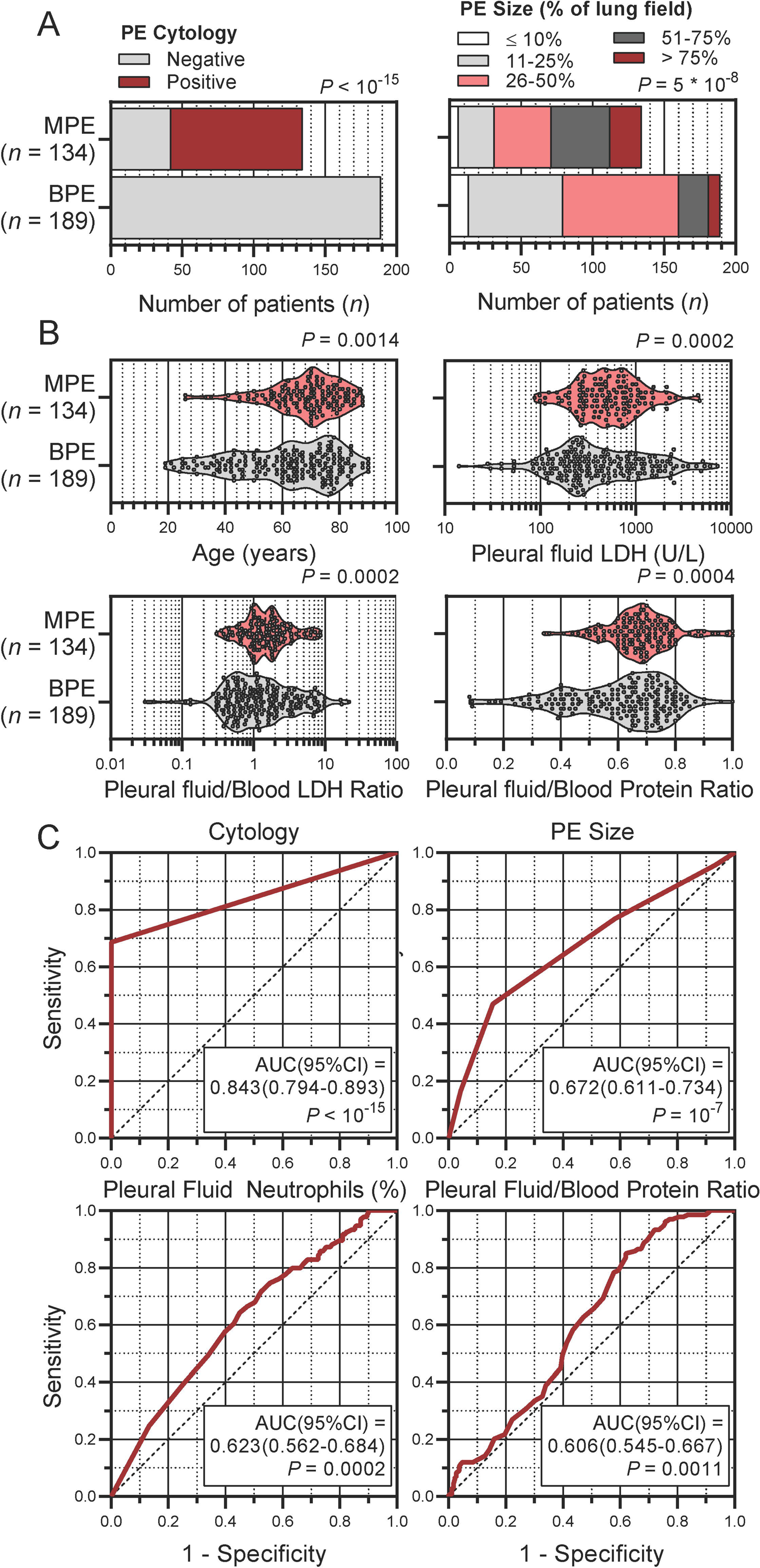
Variables significantly different between benign (BPE) and malignant (MPE) pleural effusions (PE) in the MPE detection (MAPED) study. **(A)** Frequency distributions of PE cytology and size stratified by PE diagnosis. Data are presented as patient numbers (*n*) with Fischer’s exact (left) and ^2^ (right) test probabilities (*P*). **(B)** Continuous numerical variables stratified by diagnosis. Shown are patient numbers (*n*), raw data (dots), rotated kernel density distributions (violins), median (dashed lines), quartiles (dotted lines), and Kolmogorov-Smirnov test probabilities (*P*). LDH, lactate dehydrogenase. **(C)** Shown are significant receiver-operator characteristics (curves), areas under curve (AUC) with 95% confidence intervals (95%CI), and probabilities (*P*).

**Table 1.**
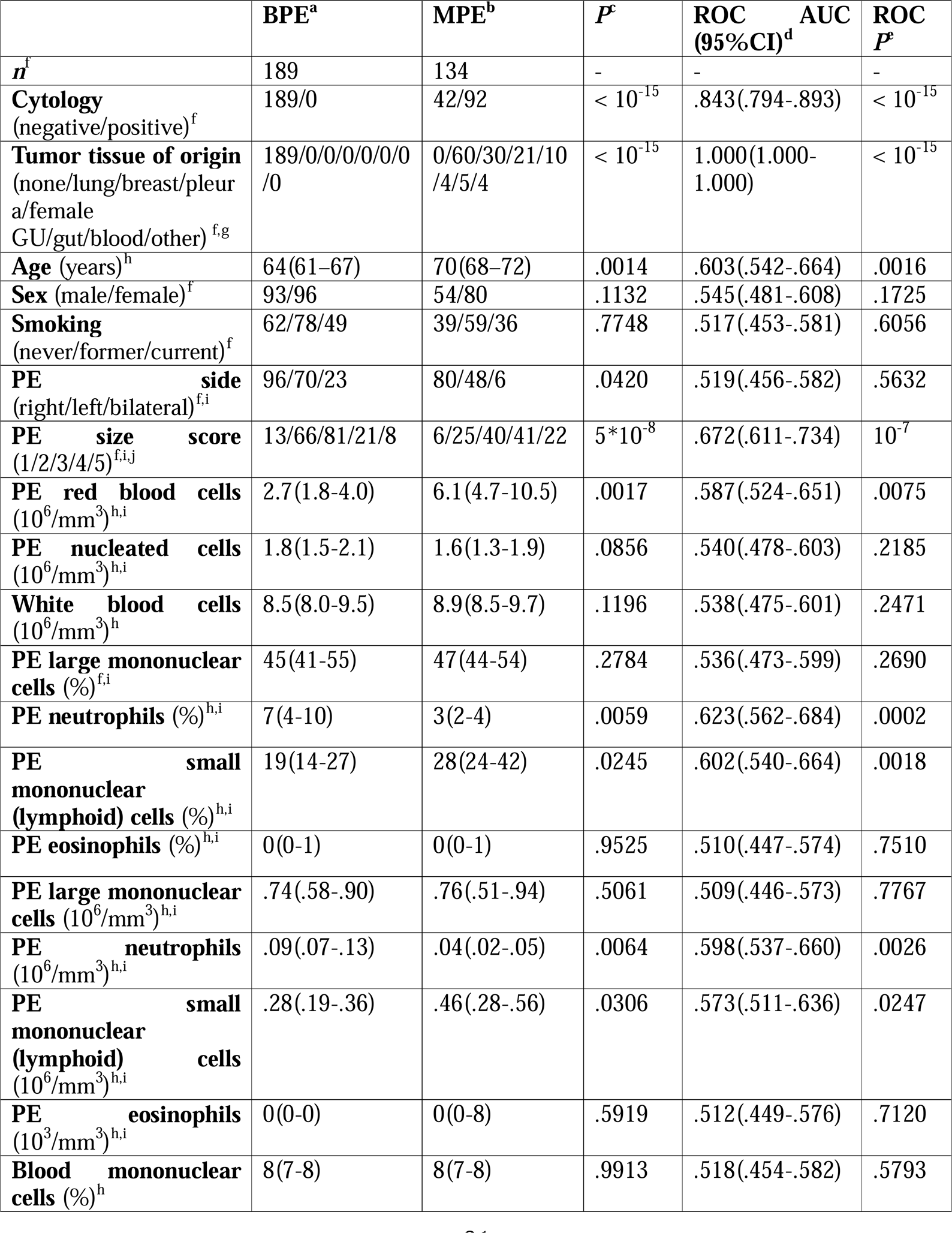
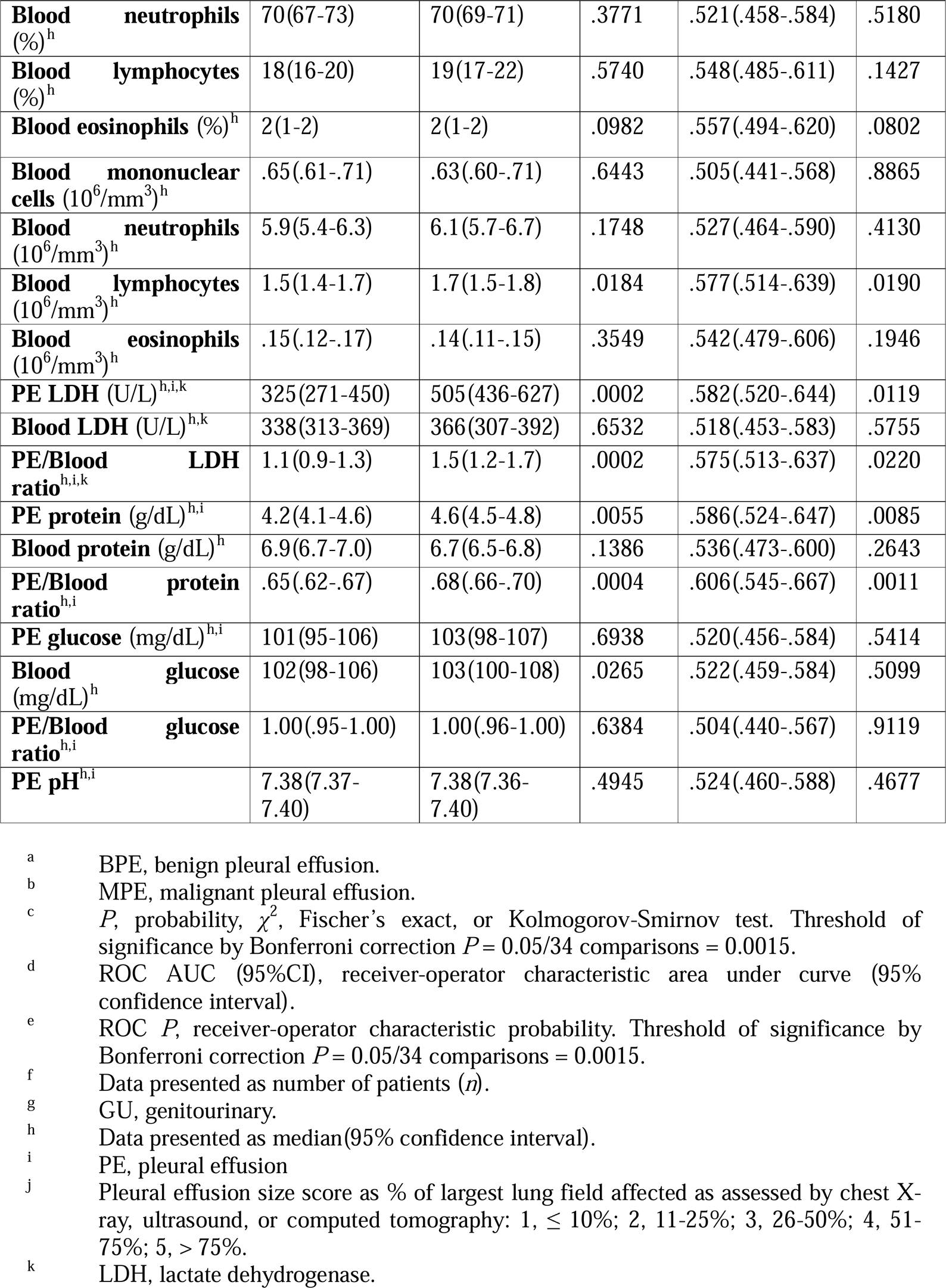
Summary of malignancy of pleural effusion detection (MAPED) study patient data at entry stratified by primary outcome.

### Machine learning identifies patient age and effusion size, neutrophil, LDH, and protein contents as the strongest predictors of underlying cancer

Subsequently, binary logistic regression and random forest analyses using MPE as target identified age, PE size, PE neutrophil count, and PE LDH and protein levels as optimal predictors of an incipient MPE diagnosis using the criteria of low probability values and mean decreased accuracy, respectively (Figure 3A). Visual inspection of partial dependency plots defined appropriate cut-offs for accurate distinction of MPE from BPE (Figure 3B). However, Euclidean distancing with agglomeration method ward.D2 (hclust function in R) was incapable of discriminating MPE apart from BPE using unsupervised hierarchical clustering (eFigure 1A). Interestingly, all five variables are routinely determined in most hospitals, except PE cytocentrifugal specimen preparation followed by PE differential cell counts, which we routinely implemented (eFigure 1B).

**Figure 3.**
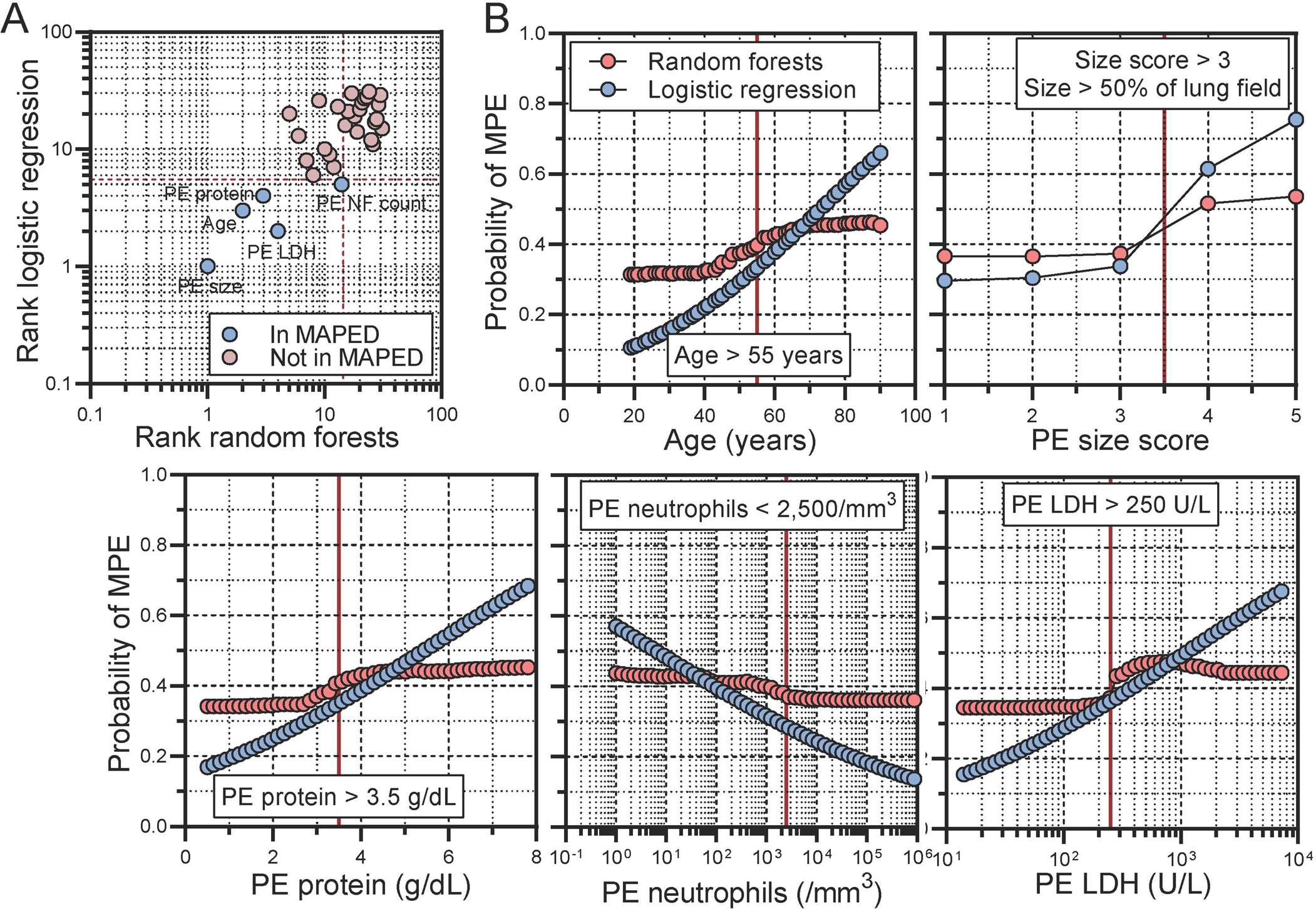
Machine learning identifies patient age, pleural effusion (PE) size, as well as PE neutrophil, protein, and lactate dehydrogenase (LDH) content as important predictors of malignant PE (MPE) in the MAPED study. **(A)** Variable importance (VIMP) plot. Data are presented as estimates (circles), cut-offs (dashed lines), and variables selected or not for inclusion into MAPED (color code). **(B)** Partial dependency plots from random forests (RF) in comparison to linear binary LR. Data are presented as probability of MPE versus benign PE (BPE) by each predictor.

### Development and validation of the MAPED score

We next incorporated age, PE size, PE neutrophil count, and PE LDH and protein levels into the MAPED score (Figure 4A). MAPED was developed and tested via resampling using the hold-out method based on 50 repeats of train/test data sets split by 70:30 ratios. To assess the predictive power of MAPED in comparison with selected logistic regression and random forest models, benchmark criteria (AUC, Brier score/skill, and AIC) were evaluated. To put the performance of MAPED into perspective, logistic regression models with all available variables or with lowest and top random forest variables were fitted to the data, and random forest models with all available and top variables were considered. Mean and standard deviations from resampling 50 test data sets for the five benchmark criteria are shown in Table 2. These analyses showed that MAPED performed equally or even superior to all random machine learning approaches employed.

**Figure 4.**
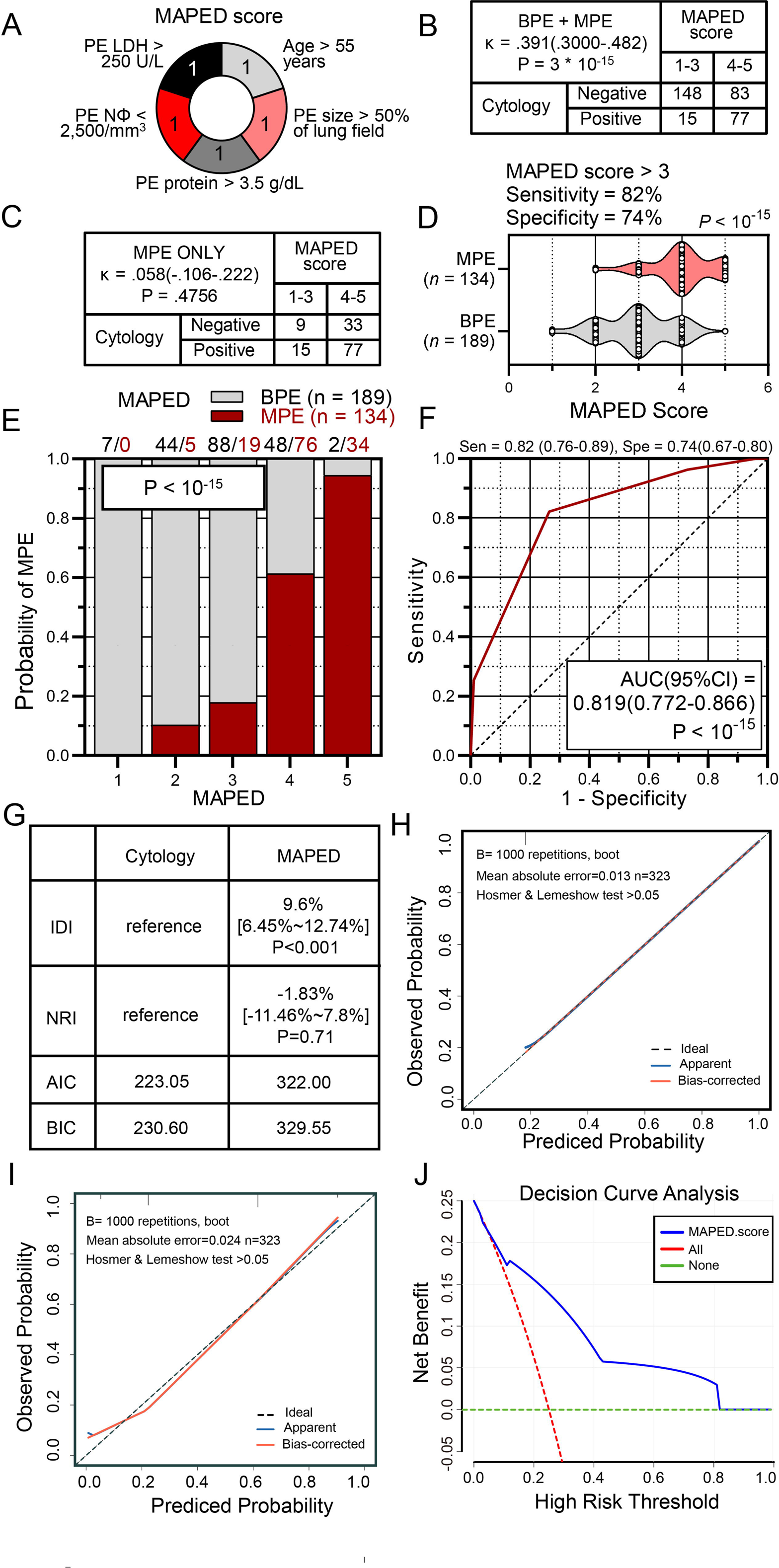
The malignant pleural effusion detection (MAPED) score and its performance in the Greek discovery. **(A)** Graphic representation of the MAPED score and its constituents. **(B, C)** Cross-tabulations of MAPED score by pleural effusion (PE) cytology results in all patients (B) and in patients with malignant PE (MPE; C). Shown are patient numbers (n), coefficients of agreement (κ), and Fischer’s exact probabilities (*P*). **(D)** Data summary of MAPED score by pleural effusion (PE) diagnosis (MPE versus benign PE, BPE). Shown are patient numbers (*n*), raw data (circles), rotated kernel density distributions (violins), median (dashed lines), quartiles (dotted lines), and Kolmogorov-Smirnov test probability (*P*). **(E, F)** Probability of MPE by MAPED score (E) and receiver-operator curve of MAPED targeting MPE diagnosis (F) in the Greek MAPED derivation cohort. Data in (E) are presented as fractions (columns), color-coded patient numbers, and probability (*P*), χ^2^ test. Data in (F) are presented as receiver-operator characteristic (curve) with area under curve (AUC), 95% confidence interval (95% CI), and probability (*P*). *n*, sample size, Sen, sensitivity; Spe, specificity. **(G)** Comparison of the discrimination and goodness of fit of two predictive models for pleural effusion (PE) in the MAPED cohort. NRI, net reclassification improvement; IDI, integrated discrimination improvement; AIC, Akaike information criterion; BIC, Bayesian Information Criterion. **(H, I)** Calibration curves of Cytology**(H)** and MAPED score **(I). (J)** Clinical net benefits in the decision curve analysis (DCA) of MAPED score.

**Table 2.** Power for MPE prediction from resampling 50 test data sets for all covariables (allCV)^a^, variables pertaining to the lowest Akaike information criterion (AIC)^b^, the five benchmark components of the malignant pleural effusion detection (MAPED) score (topCV), and the MAPED score per se by logistic regression (LR) and random forests (RF) analyses. Shown are performance measures on 70% training and 30% test data sets from 50 bootstraps. In bold font are shown the outperformers.

| Model | AUC<sup>c,d</sup> | Accuracy<sup>d</sup> | Sensitivity<sup>d</sup> | Specificity<sup>d</sup> | Brier score<sup>d</sup> | Brier skill<sup>e</sup> |
| --- | --- | --- | --- | --- | --- | --- |
| **LR, allCV** | .765±.040 | .715±.035 | .620±.073 | .784±.055 | .204±.024 | -.261 |
| **RF, allCV** | .808±.035 | .731±.036 | .543±.073 | **.866±.052** | .183±.010 | -.134 |
| **LR, low AIC** | .818±.037 | .756±.037 | .685±.071 | .807±.048 | .174±.020 | -.075 |
| **LR, topCV** | .791±.043 | .752±.040 | .635±.076 | .835±.051 | .183±.018 | -.134 |
| **RF, topCV** | .811±.037 | .727±.032 | .628±.064 | .798±.049 | .174±.018 | -0.080 |
| **LR, MAPED** | **.822±.033** | **.774±.032** | **.815±.050** | .744±.046 | **.161±.017** | **Ref<sup>f</sup>** |

<sup>a</sup> Covariables entered in raw form: smoking status, PE side, PE size, age, sex, and PE and blood differential cell percentages; and in log-transformed form: white blood and PE nucleated cell counts, PE red blood cell count, PE and blood differential cell counts, PE pH, PE and blood lactate dehydrogenase (LDH), protein, and glucose levels.
<sup>b</sup> Covariables entered in raw form: PE size, age, PE side, and PE large mononuclear, neutrophil, and eosinophil cell percentages; and in log-transformed form: PE large mononuclear cell count, PE pH, LDH, and protein levels, and blood protein levels.
<sup>c</sup> AUC, area under curve.
<sup>d</sup> Data presented as mean±SD.
<sup>e</sup> A Brier skill score has a range of $-\infty$ to 1. Negative values mean that the forecast is less accurate than a standard forecast. $< 0$ = lesser skill compared to the reference forecast. $0$ = no skill compared to the reference forecast. $1$ = perfect skill compared to the reference forecast.
<sup>f</sup> Ref, Reference.

In the MAPED cohort, the MAPED score was statistically significantly in fair agreement with cytology results when all patients were examined together, underpinning its value as a MPE-relevant end-point (Figure 4B). However, MAPED was not in agreement with cytology results when patients with MPE were examined separately (Figure 4C). This fact portrays the potential synergy of the MAPED score with cytology, since MAPED identified 33 out of 42 patients with MPE and negative cytology results (sensitivity = 79%), while it falsely incriminated only 50 patients with BPE out of the 231 with negative cytology as having cancer (specificity = 78%). These findings underscore the potential value of MAPED in the setting of negative cytology results, as well as its potential synergy with cytology in streamlining management: only 9/134(7%) patients with MPE were unidentified by both MAPED and cytology. In addition, MAPED was markedly differently distributed in patients with BPE and MPE, and identified MPE with AUC = 82% in the derivation cohort (Figures 4D-4F). To further determine how well the MAPED model reclassifies patients into MPE and BPE groups, we calculated IDI and NRI parameters and compared them with the Cytology model. The MAPED model significantly improved the classification ability compared to Cytology (IDI _MAPED_ = 9.6%, IDI _Cytology = reference_, p < 0.001, Figures 4G). There was no difference in NRI between the above two models, using cutoffs of 0%-30% (low risk), 30%-60% (moderate risk), and 60%-100% (high risk). (Figures 4G). These results indicate that MAPED and Cytology have comparable discriminatory power. Since calibration reflects the extent to which the model correctly assesses absolute events or risks^(14)^, we calculated AIC and BIC, which revealed the following:

AIC = MAPED (322.00): Cytology (223.05) and BIC = 329.55:230.60 (Figures 4G). These results suggest a slightly better calibration in the Cytology model compared to the model in MAPED. The calibration curves between predicted and observed values for the Cytology model fit the predicted probabilities along the x-axis well and slightly better than the MAPED model (Figures 4H-I). However, the Hosmer & Lemeshow test of p>0.05 indicates that the MAPED model is well-calibrated overall (Figures 4I). To assess the clinical significance of MAPED, we performed the DCA, which showed that the MAPED model had a net clinical benefit of significant within a high-risk threshold probability range of 0.1-0.8 (Figure 4J). Subsequently, the MAPED cohort was randomly divided into two cohorts according to split by 70:30 ratios. In the cohort 1 dataset (70%), the analysis was significantly consistent with the entire dataset (eFigure 2), and the discriminative power of the MAPED curve was consistent with that shown in the entire dataset (MAPED vs. Cytology; AUC = 0.829 vs. 0.832, eFigure 2A-B). Calibration curves for both models were similar to the entire dataset and had a good fit (eFigure 2C-D). DCA analysis also showed that the MAPED model had a significant net clinical benefit within the high-risk threshold probability range of 0.1-0.8 (eFigure 2E). In the cohort two dataset (30%), the analysis generally showed consistent results from the entire dataset (eFigure 3).

We finally determined the accuracy of MAPED in the ORPB, one of the few cohorts where PE size was determined and where PE neutrophil data are available, although in a different format compared with MAPED (neutrophil versus lymphocyte predominance is determined in most hospitals worldwide as compared with our quantitative data). Despite these discrepancies, MAPED performed reasonably well in 238 (125 with BPE and 113 with MPE) ORPB patients, correctly predicting MPE with AUC = 72% (Figures 5A and 5B; Table 3). The calibration curve of the MAPED model and the Hosmer & Lemeshow test with p>0.05 indicated that the MAPED model was generally well calibrated (Figure 5C). The results of the DCA indicate that the MAPED model has a significant net clinical benefit in the high risk threshold probability range of 0.1 to 0.6 (Figure 5D).

**Figure 5.**
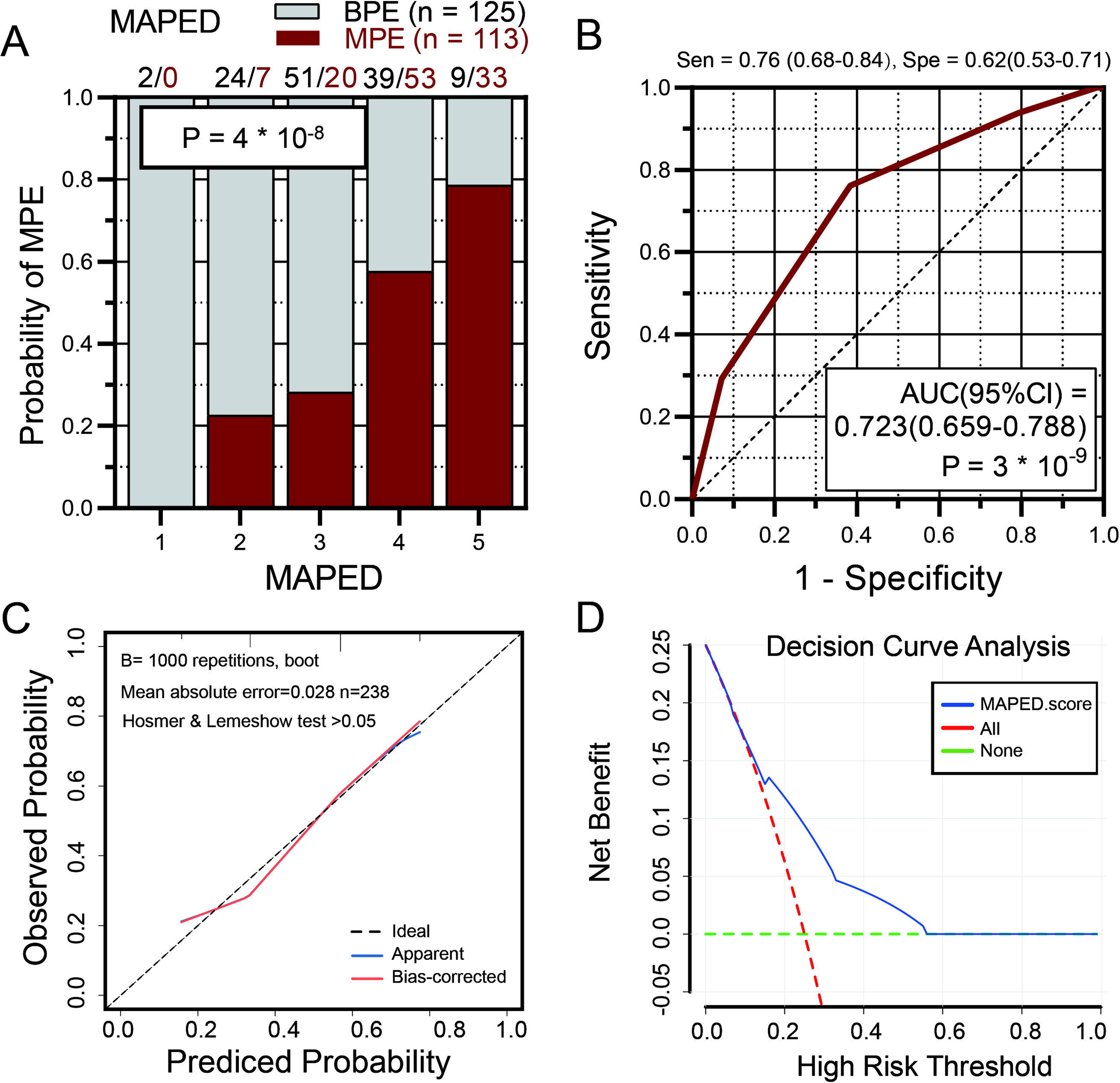
The malignant pleural effusion detection (MAPED) score and its performance in the UK validation cohort. **(A, B)** Probability of MPE by MAPED score **(A)** and receiver-operator curve of MAPED targeting MPE diagnosis **(B)** in the UK Oxford Radcliffe Pleural Biobank (ORPB) validation cohort. Data in **(A)** are presented as fractions (columns), color-coded patient numbers, and probability (*P*), ^2^ test. Data in **(B)** are presented as receiver-operator characteristic (curve) with area under curve (AUC), 95% confidence interval (95% CI), and probability (*P*). *n*, sample size. Sen, sensitivity; Spe, specificity. **(C)** Calibration curves of MAPED score. **(D)** Clinical net benefits in the decision curve analysis (DCA) of MAPED score.

**Table 3.**
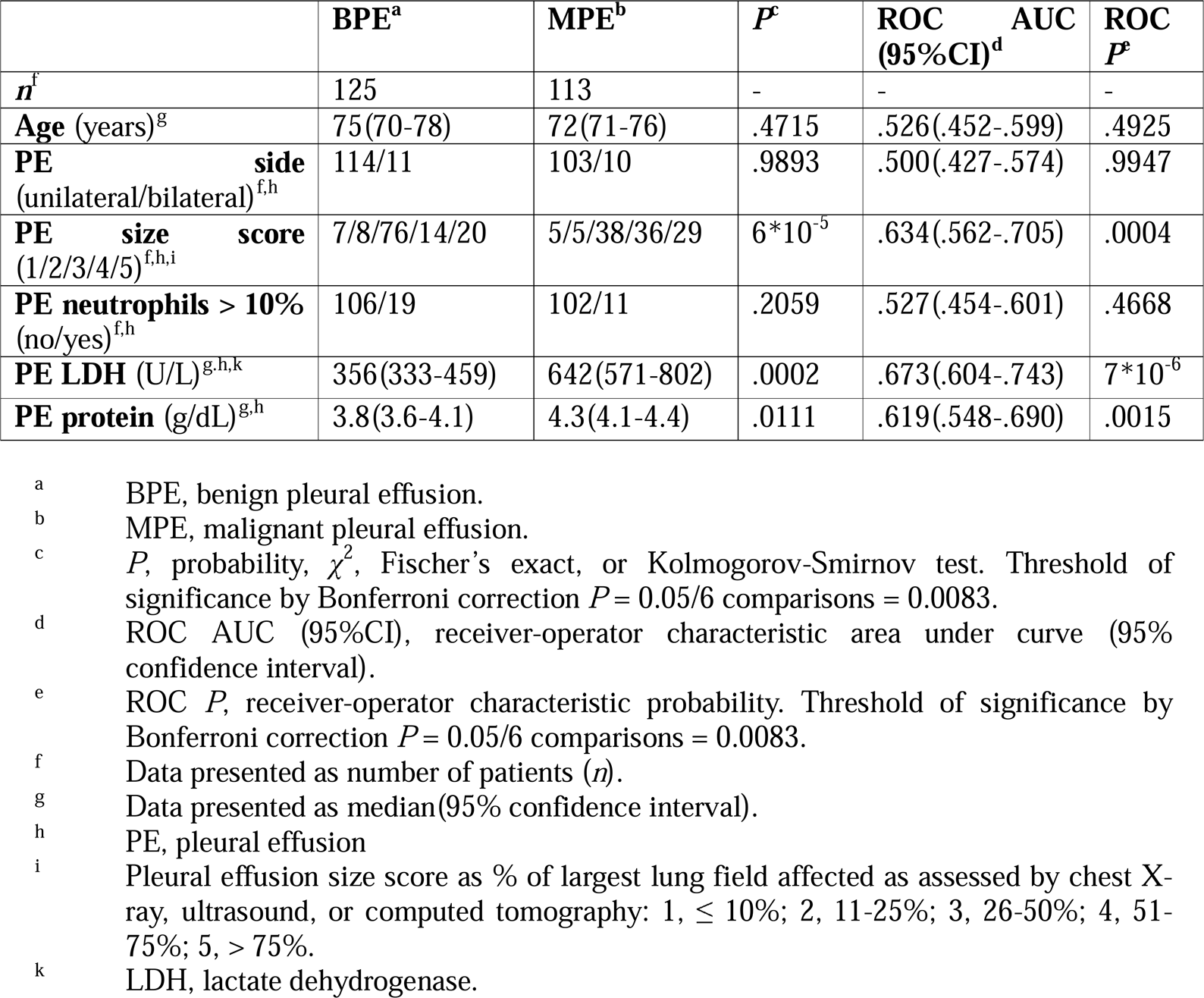
Summary of Oxford Radcliffe Pleural Biobank (ORPB) study patient data stratified by primary outcome.

|  | **BPE<sup>a</sup>** | **MPE<sup>b</sup>** | **P<sup>c</sup>** | **ROC AUC (95%CI)<sup>d</sup>** | **ROC P<sup>e</sup>** |
| --- | --- | --- | --- | --- | --- |
| **n<sup>f</sup>** | 125 | 113 | - | - | - |
| **Age (years)<sup>g</sup>** | 75(70-78) | 72(71-76) | .4715 | .526(.452-.599) | .4925 |
| **PE side (unilateral/bilateral)<sup>f,h</sup>** | 114/11 | 103/10 | .9893 | .500(.427-.574) | .9947 |
| **PE size score (1/2/3/4/5)<sup>f,h,i</sup>** | 7/8/76/14/20 | 5/5/38/36/29 | 6*10<sup>-5</sup> | .634(.562-.705) | .0004 |
| **PE neutrophils > 10% (no/yes)<sup>f,h</sup>** | 106/19 | 102/11 | .2059 | .527(.454-.601) | .4668 |
| **PE LDH (U/L)<sup>g,h,k</sup>** | 356(333-459) | 642(571-802) | .0002 | .673(.604-.743) | 7*10<sup>-6</sup> |
| **PE protein (g/dL)<sup>g,h</sup>** | 3.8(3.6-4.1) | 4.3(4.1-4.4) | .0111 | .619(.548-.690) | .0015 |

<sup>a</sup> BPE, benign pleural effusion.
<sup>b</sup> MPE, malignant pleural effusion.
<sup>c</sup> P, probability, $\chi^2$ , Fischer's exact, or Kolmogorov-Smirnov test. Threshold of significance by Bonferroni correction $P = 0.05/6$ comparisons = 0.0083.
<sup>d</sup> ROC AUC (95%CI), receiver-operator characteristic area under curve (95% confidence interval).
<sup>e</sup> ROC P, receiver-operator characteristic probability. Threshold of significance by Bonferroni correction $P = 0.05/6$ comparisons = 0.0083.
<sup>f</sup> Data presented as number of patients (n).
<sup>g</sup> Data presented as median(95% confidence interval).
<sup>h</sup> PE, pleural effusion
<sup>i</sup> Pleural effusion size score as % of largest lung field affected as assessed by chest X-ray, ultrasound, or computed tomography: 1, $\leq 10\%$ ; 2, 11-25%; 3, 26-50%; 4, 51-75%; 5, $> 75\%$ .
<sup>k</sup> LDH, lactate dehydrogenase.

The nomogram was constructed to predict patient risk scores to make the MAPED score more convenient for physicians to use in clinical practice (Figure 6). Taking together both cohorts, a MAPED score of > 3 points was determined in *n* = 294 patients and correctly identified 196 of 247 MPE, yielding a sensitivity of 79%, while falsely incriminating 98 of 314 BPE, for a specificity of 69%.

**Figure 6.**
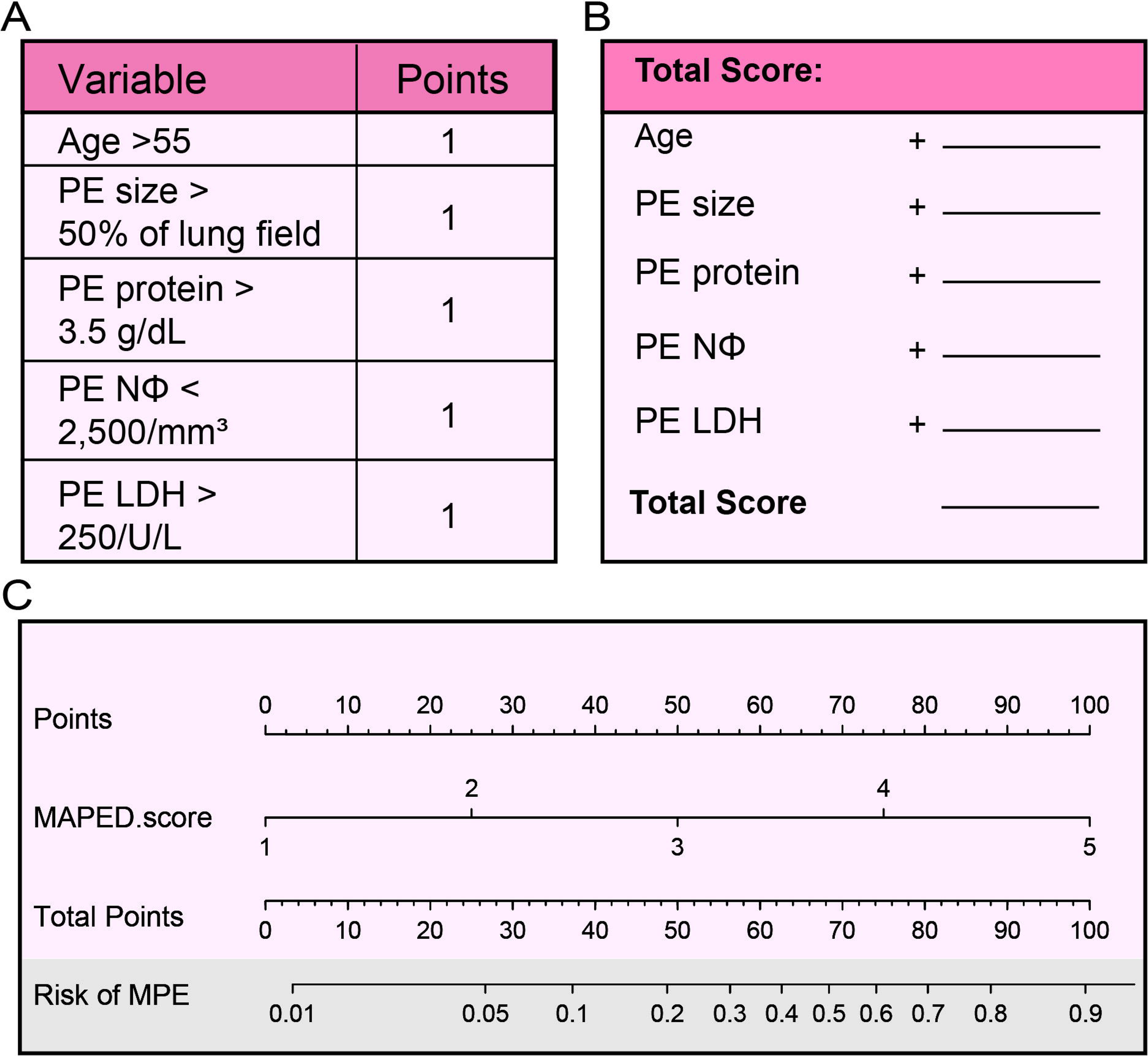
Brief score table and diagnostic nomogram for distinguishing MPE from BPE. **(A)** Points assigned to the individual variable. **(B)** Calculation of the total risk score of the patient. **(C)** Diagnostic nomogram for identifying BPE from BPE.

## DISCUSSION

MAPED addresses an unmet clinical need: to determine the likelihood of a MPE during initial patient work-up. We show that five variables combined into the MAPED score can predict MPE in 82% of cases in the MAPED derivation cohort, with discrimination and calibration performing almost equally to cytology. Importantly, the MAPED score also predicted MPE in 72% of the ORPB validation cohort. We also show that MAPED performs differently than cytology in patients with MPE, and can supplement cytologic investigation in streamlining management. For example, the current clinical practice of repeat cytology and/or tissue biopsy in cytology-negative patients with PE could be improved by prioritizing the 33 MAPED-high cytology-negative patients with MPE and the 50 MAPED-high cytology-negative patients with BPE for intensive cytology (cytospins/cellblocks/immunocytochemistry) and pleural tissue biopsy (*n* = 83 total patients prioritized) over our 231 total patients investigated. This would come at a cost of only 9 patients (< 3%) with MPE that were missed by both cytology and MAPED, thereby decreasing time to and cost of diagnosis.

The accuracy of MAPED is satisfactory given its simplicity. Another effort to build a similar score included patients with uncertain diagnoses, had multiple primary end-points, and lacked external validation^(15)^. A chest computed tomography (CT)-based score derived from 343 prospectively enrolled patients with PE achieved AUC = 0.919 in discriminating MPE from BPE ^(16)^. However, contrast-enhancement and scan reading by two blinded experienced radiologists was required, increasing risk, cost, and time. Despite the above, inter-observer agreement was only 0.55–0.94. To scan our 323 discovery and 238 validation patients assuming a cost of € 200/scan and 0.5 hour physician time required for scan interpretation would cost € 112,200 and 280.5 radiologist hours. We used routine bedside tests to build MAPED, which performs only slightly inferior to the above-referenced CT score, at zero additional cost and physician time.

The MAPED score components are worth mentioning here. Age is linked with cancer development ^(17)^, but its value in prospectively differentiating MPE from BPE has never been exploited, as most studies did not detect age differences between patients with MPE and BPE ^(1,2,4,18)^. We did, and although the mean age difference between our patients with BPE and MPE was small (six years), an age cut-off of > 55 years alone could discriminate MPE from BPE with AUC = 0.603. This was not the case in the ORPB validation set, where an age cut-off of > 55 years produced an AUC = 0.526 (*P* = 0.4925). Notwithstanding population (https://knoema.com/atlas/topics/Demographics/Age/Median-age-of-population?origin=knoema.de; accessed on 08.04.2021) and healthcare accessibility (https://ourworldindata.org/grapher/healthcare-access-and-quality-index; accessed on 08.04.2021) differences between Greece (2015 mean age = 43.4 years and healthcare access & quality index = 87) and the UK (2015 mean age = 40.0 years and healthcare access & quality index = 84.6) that can explain this discrepancy and may necessitate different age cut-offs in different countries, we chose to develop a generally applicable MAPED score and applied it to ORPB patients.

Relative neutrophil predominance in pleural fluid is also a well-known hallmark of infectious BPE due to common pathogens ^(3)^, but has not been used as a negative marker of PE malignancy. Interestingly, one important study identified blood neutrophil-to-lymphocyte count ratio as an important determinant of the survival of patients with MPE ^(2)^. However, the use of relative pleural neutrophil abundance to rule out MPE is hampered by the common practice of not accurately counting PE cells by most hospitals in the US and Europe. We overcame this by establishing PE differential counts as routine practice for the purposes of MAPED. The effort was worth spent, since neutrophil counts < 2,500/mm^3^ produced a statistically significant AUC = 0.598 (*P* = 0.0026) in MAPED. Again, this was not the case in ORPB, where pleural neutrophil paucity ≤ 10% produced a non-significant AUC = 0.527 (*P* = 0.4668). However, neutrophil paucity in ORPB was defined semi-quantitatively on PE smears and not quantitatively on PE cytospins as in MAPED.

Unlike the aforementioned predictors of MPE that failed to perform well in the ORPB validation cohort, PE size > 50% of the most affected lung field, PE protein levels > 3.5 g/dL, and PE LDH levels > 250 U/L did perform excellently in both MAPED and ORPB. In specific, only 27% of BPE but an astonishing 58% of MPE fulfilled the > 50% of the most affected lung field size criterion in ORPB (*P* = 6*10^−5^; χ^2^ test) compared with 15% and 47% in MAPED (*P* = 5*10^−8^; χ^2^ test), respectively. In addition, 60% of BPE and 78% of MPE fulfilled the protein criterion in ORPB (*P* = .0030; χ^2^ test) compared with 68% and 87% in MAPED, respectively (*P* = 7*10^−5^; χ^2^ test); and 65% of BPE and 84% of MPE fulfilled the LDH criterion in ORPB (*P* = .0007; χ^2^ test) compared with 63% and 86% in MAPED, respectively (*P* = 6*10^−6^; χ^2^ test); with the similar results probably owing to more uniform methods of measurement, since pleural protein and LDH levels are established Light’s criteria ^(3, 13)^. The size, protein, and LDH criteria also produced significant AUC values in ORPB, comparable to those achieved in MAPED (Tables 1 and 2). Although it is well established that MPE pathogenesis includes increased vascular permeability leading to protein-rich exudate ^(19)^, pleural fluid-to-blood protein ratio is an exudate criterion according to Light ^(3,13)^, and protein measurements are routine in contemporary hospitals, PE protein levels have never been exploited to diagnose MPE. To this end, PE LDH > 1500 U/L was recently proposed as a poor prognosis marker for MPE (2), and high MPE protein levels were found in a previous study ^(15)^, rendering our findings plausible. Massive PE have rarely been studied separately, although they are common with both BPE and MPE ^(20, 21)^. In the largest study looking at PE size, Porcel *et al*. classified 535 patients with BPE and 231 with MPE into three size categories based on posterior-anterior chest X-rays: non-large PE was defined as occupying less than two thirds of the lung field, large as occupying more than that, and massive as occupying the whole lung field ^(20)^. Interestingly and in accord with our results, the authors found that 24% of non-large, 49% of large, and 59% of massive PE were malignant (*P* < 10^−5^; χ^2^ test), but this pearl has never been used to estimate MPE risk.

The present study has limitations. First, the general applicability of MAPED is hampered by population, measurement, and practice differences between countries, as well as by divergent prevalence of specific causes of PE. However, MAPED was only loosely fit to the derivation cohort by avoiding weighing, and hence withstood testing in such a remote setting. In addition, the relative prevalence of MPE in MAPED (40% of all PE) was similar to most other published studies from Europe and North America that report values from 30– 54%^(1,15,21)^. A second limitation is chest X-ray interpretation, the only non-standardized measure included in MAPED. However, estimating the percentage of a lung field occupied by a PE is an easy task. Third, MAPED cannot be used in outpatients, and patients with previous PE or cancer, by design. Finally, MAPED was not designed to detect MPE that are diagnosed many years past initial PE presentation ^(22^).

In conclusion, We develop a novel scoring system and translate it into a nomogram scoring system so clinicians can predict the probability of MPE and BPE. The MAPED score is based on various clinical and laboratory indicators available in most hospitals and even in primary care. Importantly, the MAPED score correctly classified 412 of 561 (73.44%) pleural effusions examined in two countries, at no additional risk to patients, cost to healthcare systems, and time spent to caring physicians. Pending further validation, MAPED may contribute to improvements in patient management and research design, since it alters the likelihood of MPE at admission as a rule out or rule in score.

**Author Contributions:** JJ, AM, AV, IP, ASL, MI, ACK, IL, KK, KS and RL performed and analyzed the clinical study form Greece cohort; JJ, RA, NIK, NMR, and IP performed and analyzed the clinical study form Oxford cohort; JCK did regression and random forest analyses; JJ, GTS conceived the main idea and steered the study, performed data analyses and graphic design, and wrote the paper. GTS is the guarantor of the content of the manuscript, including data and analysis. All authors contributed to the conception, design, acquisition, analysis, interpretation, drafting, and revising the work. All authors approved the version to be published and agree to be accountable for the work.

**Funding:** This work was supported by European Research Council 2010 Starting Independent Investigator (#260524) and 2015 Proof of Concept (#679345) grants, the Graduate College (Graduiertenkolleg, GRK) #2338 of the German Research Society (Deutsche Forschungsgemeinschaft, DFG), the target validation project for pharmaceutical development ALTERNATIVE of the German Ministry for Education and Research (Bundesministerium für Bildung und Forschung, BMBF), and a Translational Research Grant by the German Center for Lung Research (Deutsches Zentrum für Lungenforschung, DZL) (all to GTS). The study sponsors had no role in study design, data collection, analysis, and interpretation, and in writing and submitting the paper for publication.

**Conflict of interest:** I.P. is employed as a Global Clinical Head by Astra Zeneca Pharmaceutical in a field that is non-related with the publication. The remaining authors have declared that no conflict of interest exists.

## Data Availability

All data are available by the corresponding author upon request.

