## supplementary material for "Clinical identification of malignant pleural effusions"

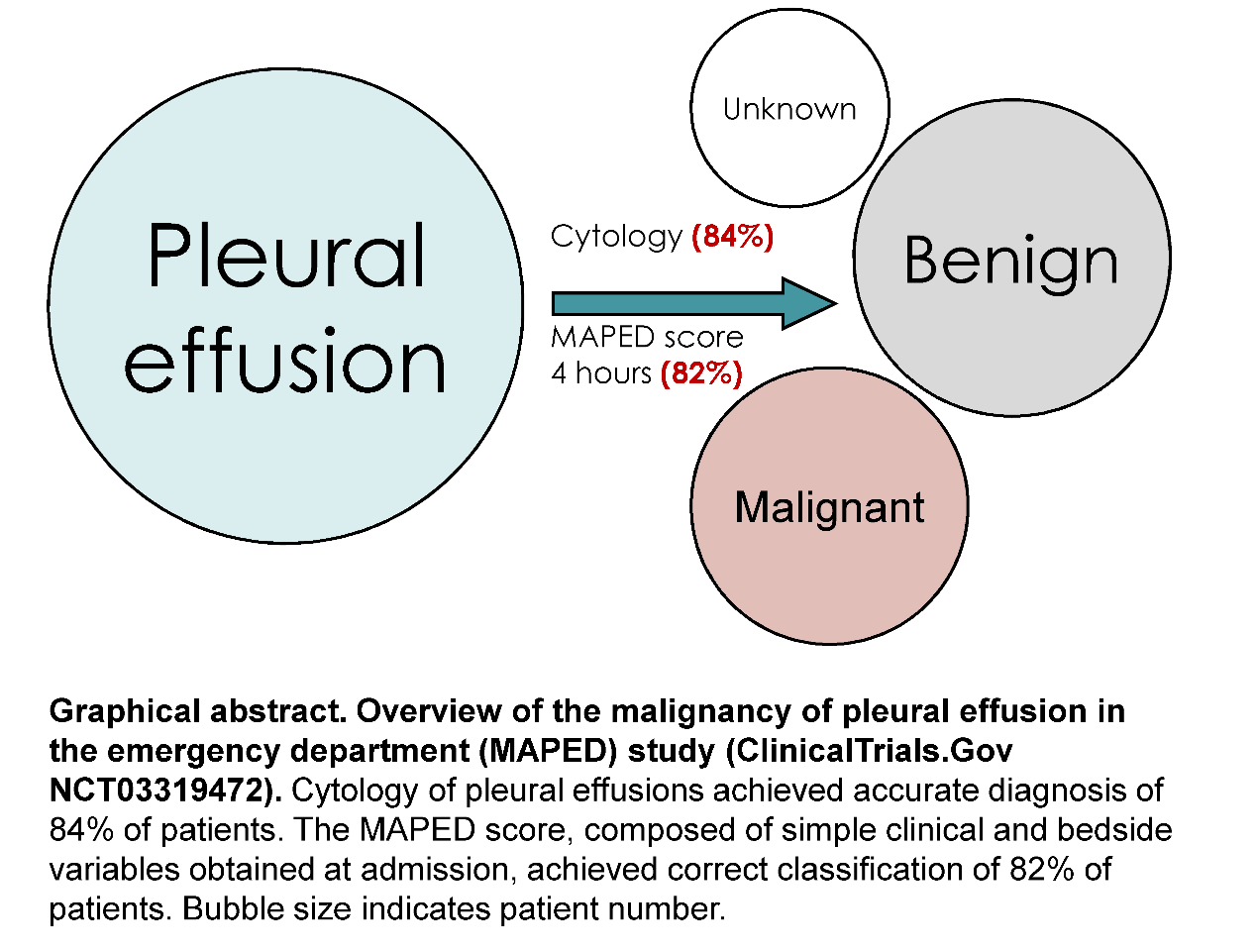

**Graphical abstract**

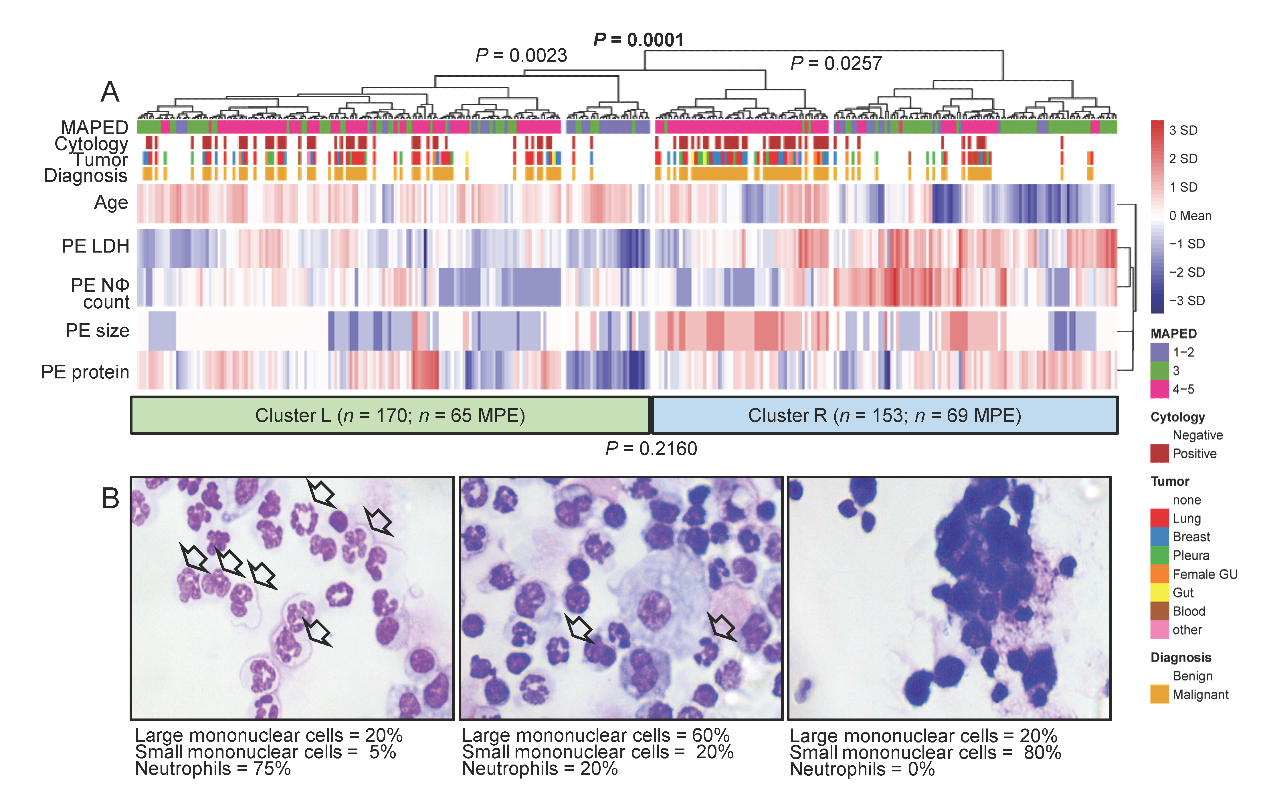

**eFigure 1. Unsupervised hierarchical clustering alone cannot distinguish benign from malignant pleural effusions (PE).** **(A)** Heatmap shows unsupervised hierarchical clustering of *n* = 323 patients by patient age, PE size, as well as PE neutrophil (NΦ), protein, and lactate dehydrogenase (LDH) content. Each row represents one marker and each column one patient. *P*, probability, top: Euclidean distance between clusters (the smallest P value was used to separate the two major clusters), bottom: hypergeometric test for enrichment of MPE in any of the two clusters L (left) and R (right). **(B)** Representative PE cytocentrifugal specimens (cytospins) with differential cell counts. Arrows indicate neutrophils.

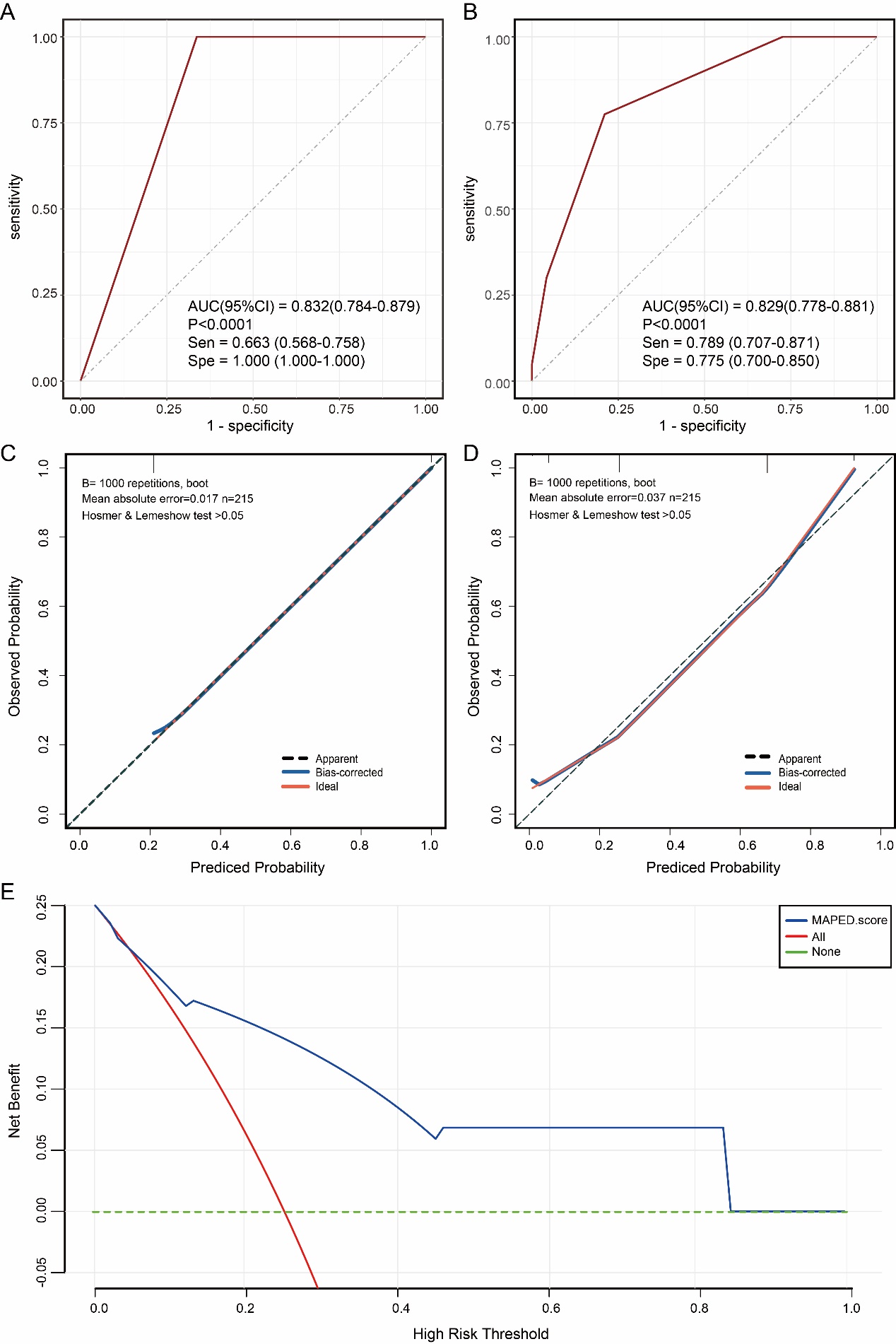

**eFigure 2. The malignant pleural effusion detection (MAPED) score and its performance in the cohort 1 dataset (70% of MAPED cohort).**

**(A-B)** Receiver operating characteristic (ROC) curves and area under the curve (AUC) in the Cytology**(A)** and MAPED score**(B)** models. Sen，sensitivity; Spe, specificity. **(C,D)** Calibration curves of Cytology**(C)** and MAPED score **(D)**.  **(E)** Clinical net benefits in the decision curve analysis (DCA) of MAPED score.

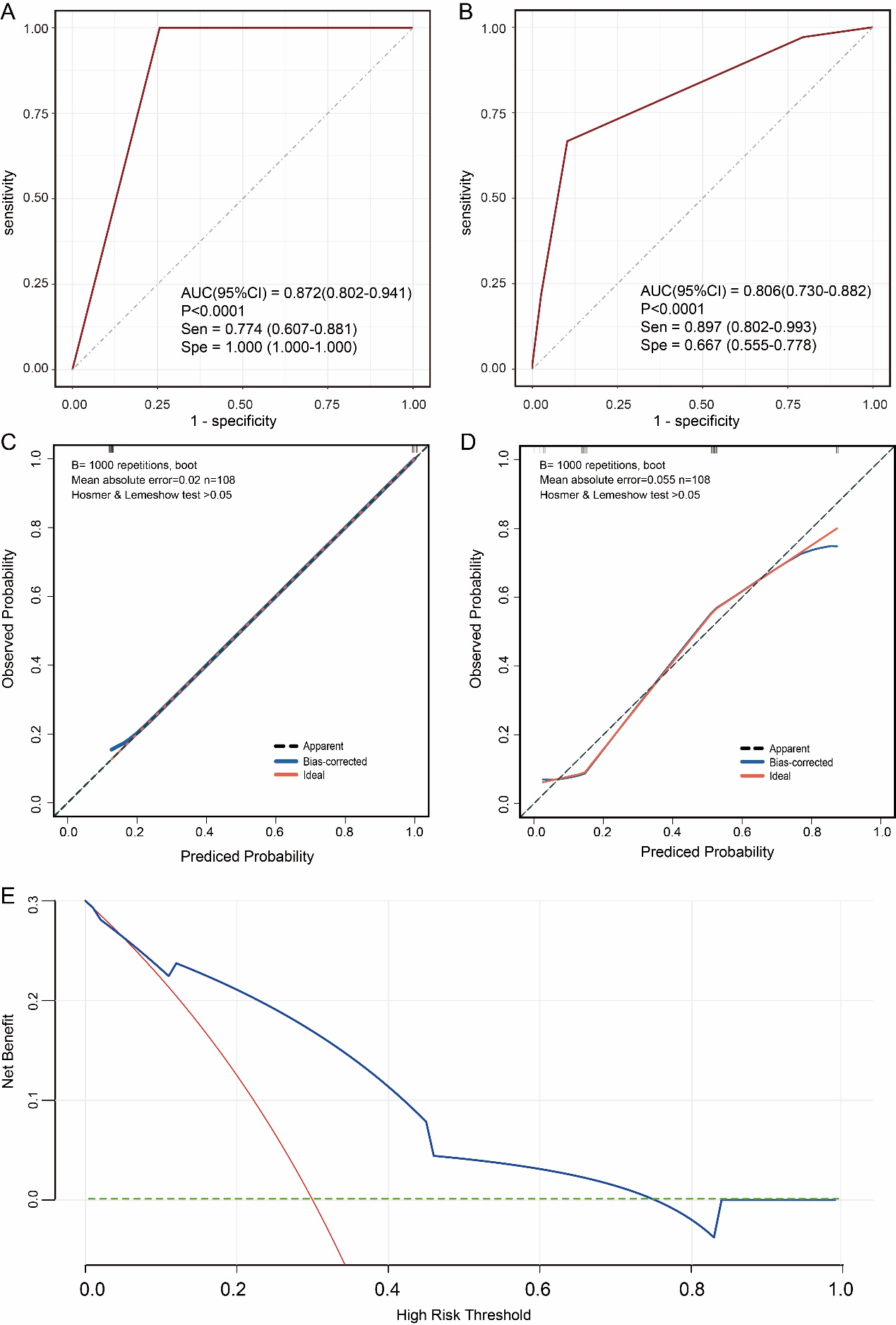

**eFigure 3. The malignant pleural effusion detection (MAPED) score and its performance in the cohort 2 dataset (30% of MAPED cohort).**

**(A-B)** Receiver operating characteristic (ROC) curves and area under the curve (AUC) in the Cytology**(A)** and MAPED score**(B)** models. Sen，sensitivity; Spe, specificity. **(C,D)** Calibration curves of Cytology**(C)** and MAPED score **(D)**. **(E)** Clinical net benefits in the decision curve analysis (DCA) of MAPED score.
